# Phase II study of nivolumab and ipilimumab for treatment of metastatic/recurrent adenoid cystic carcinoma (ACC) of all anatomic sites of origin and other malignant salivary gland tumors

**DOI:** 10.1101/2024.04.29.24306581

**Authors:** Young Kwang Chae, Richard Duan, Liam Il-Young Chung, Youjin Oh, Borislav Alexiev, Sangwon Shin, Sukjun Kim, Irene Helenowski, Maria Matsangou, Victoria Villaflor, Devalingam Mahalingam

## Abstract

**Objective:** Dual checkpoint inhibitor therapy with nivolumab and ipilimumab has been FDA-approved for a number of cancer sites. However, their role in the treatment of ACC and other salivary gland carcinomas is not well established.

**Methods and analysis:** We performed a Simon’s two-stage prospective single-institution phase II clinical trial of nivolumab with ipilimumab. Two cohorts were analyzed: patients with metastatic/recurrent ACC and patients with other salivary gland cancers. The primary endpoint was median progression-free survival (PFS); secondary endpoints were overall response rate (ORR), overall survival (OS), and toxicity.

**Results:** Patients with ACC (n=19) and other salivary gland carcinomas (total n=5) were enrolled. The patients with ACC had median OS 30.0 months (95% CI 15.3-NR months), median PFS 8.3 months (95% CI 5.5-30.0 months), and disease control rate (DCR) 53% (10/19). The ORR in the ACC group was 5% (CR 0%, n=0; confirmed PR 5%, n=1), with one patient having continued stable disease at the time of trial conclusion. The patients with other salivary gland cancers had median OS 10.4 months (95% CI 6.21-NR months), median PFS 6.21 months (95% CI 2.83-NR months), and DCR 40% (2/5). The ORR in this cohort was 0%.

**Conclusion:** In patients with recurrent or metastatic ACC and other salivary gland neoplasms, combination nivolumab with ipilimumab resulted in moderate disease control. Further studies are warranted to validate our findings.

Trial number: NCT03146650

## Introduction

Adenoid cystic carcinoma (ACC) is a subset of adenocarcinomas that primarily occurs in the major and minor salivary glands. It affects 1,200 individuals in the US each year and comprises 10% of all cases of salivary gland tumors[1,2]. The standard of treatment for localized ACC cases is surgical excision followed by radiotherapy[3]; however, despite treatment, ACC cases are characterized by perineural invasion and hematologic metastasis, most notably to the lungs[4]. This leads to frequent local and distant recurrences, resulting in a 10-year survival rate of 61%[4].

Despite the persistence of ACC, it has an indolent course at the beginning. This makes it difficult to study clinical responses to chemotherapies, which have yet to demonstrate a positive effect on patient survival[1,2]. Due to the ineffectiveness of cytotoxic chemotherapies[5], more research is needed for therapies that target different molecular pathologies of ACC. MYB translocations, c-kit overexpression, EGFR overexpression, histone deacetylase aberrant activity, NOTCH1 mutations, and immunotherapy susceptibility are all examples among other attributes[6].

Many experimental therapies thus far have undergone trials to attempt to exploit the mutations and biomarkers specific for ACC. The MYB transcription factor, the most well-studied accessory of ACC oncogenesis, is mutated in about 70% of ACC patients[6], and a number of pharmaceutical targets along its signaling pathway have been identified. However, efforts to target this pathway have had little success, although a number of trials are still underway to further investigate this pathway as a therapeutic target[7]. An EGFR-targeting drug cetuximab, when combined with cisplatin, showed an overall response rate of 43%[5]. However, kinase inhibitors (e.g. c-kit inhibitors dasatinib and imatinib, multikinase inhibitors dovitinib, axitinib, and sorafenib) showed little or no clinical response with the exception of rivoceranib, a VEGF receptor tyrosine kinase inhibitor that had 14 and 17 percent ORR rates in patients with and without prior systemic treatment respectively[8,9].

Vorinostat and chidamide, two histone deacetylase inhibitors, were shown to induce limited responses in ACC patients, even when the latter was used together with cisplatin[8,10]. The NOTCH signaling pathway has been a target of a number of recent Phase I trials: a NOTCH1 antibody bronticuzumab, an upstream inhibitor of the NOTCH pathway CB-103, and an inhibitor of NOTCH signaling AL101[7]. However, these agents all showed minimal responses among patients in their respective trials, and research into targeted therapies remains crucial for improving the outcomes of ACC patients.

PD-1 and CTLA-4 are two cell surface receptors that serve as inhibitory checkpoints for activated T-cells when bound. Two immune checkpoint inhibitors, nivolumab and ipilimumab, target PD-1 and CTLA-4 respectively to prevent abrogation of T cell activation[11]. Although a trial of nivolumab alone did not show much efficacy with overall response rates below 10%[12], the nivolumab/ipilimumab combination holds some promise in the treatment of ACC. This combination of immune checkpoint inhibitors has already been FDA-approved for the treatment of metastatic or recurrent NSCLC, melanoma, renal cell carcinoma, hepatocellular carcinoma, and microsatellite insatiable-high/mismatch repair deficient colorectal cancers[11]. Furthermore, past nivolumab/ipilimumab trials for adenoid cystic salivary and other salivary neoplasms have shown dramatic tumor regression changes in the patients who did exhibit positive responses[13,14]. The objective of this study is to further elucidate the efficacy of the nivolumab/ipilimumab combination in the treatment of ACC and elucidate differences in responses between immunogenic and non-immunogenic cases of ACC.

## Methods

### Patients

Eligibility criteria include a confirmed diagnosis of metastatic/recurrent ACC from any anatomical origin or non-ACC neoplasms of a salivary gland; non-candidacy for curative surgical or radiation therapy; measurable disease in accordance with RECIST v1.1; at least 18 years of age; an Eastern Cooperative Oncology Group status of 0-2 with 3 allowed if directly secondary to ACC; adequate bone marrow and organ function; no investigational or chemotherapies administered within 28 days or radiation within 7 days of study treatment; and no history of autoimmune disease that may affect organ function or require immunosuppressive treatment. Study approval was given by the institutional IRB in accordance with federally mandated regulations. Patient informed consent was individually obtained in written form, and patients were not involved in the design and implementation of this study.

### Intervention

Patients were treated with nivolumab at 240 mg intravenously every 2 weeks for 16 weeks, then starting on Cycle 2 Day 29 given at 480 mg IV every 4 weeks. Ipilimumab was administered at 1 mg/kg intravenously every 6 weeks. Patients were assessed for response every 12 weeks (+/- 7 days). Treatment was provided until disease progression, intercurrent illnesses preventing further treatment, treatment delays greater than 42 days, patient withdrawal, or unacceptable toxicities and adverse events. All patients who received at least one dose of nivolumab and had a radiologic evaluation of disease were evaluated for efficacy endpoints. Adverse events were graded using version 4.0 of the National Cancer Institute’s Common Terminology Criteria for Adverse Events and followed up to 30 days after treatment cessation or initiation of a new treatment.

### Endpoints

A Simon two stage optimum Phase II design was used for this trial with main outcomes of PFS at 6 and 12 months (supplemental methods). Recruitment in the first stage was planned to halt after 15 subjects joined. Accrual to the next stage was contingent upon less than 7 of the 15 patients withdrawing from the study under reasons unrelated to withdrawal of consent within a timespan of 24 weeks after drug administration. In the actual trial, 10/15 patients remained after 24 weeks. Thus, the trial entered the second stage, where 43 patients were to be recruited. Due to funding constraints, trial enrollment concluded after 9 additional patients were accrued.

Patients with recurrent/metastatic salivary gland tumors were included as an exploratory group with separate statistical analysis and safety analysis. The study was designed for 10 patients to initially be enrolled, with accrual of an additional 10 patients contingent on 2 or less of the initial 10 non-ACC patients withdrawing after 24 weeks of treatment. Only 5 patients with recurrent/metastatic salivary gland cancers were ultimately enrolled due to funding constraints.

The primary end point of the study was median progression-free survival (PFS) and PFS at 6 months based on radiographic assessment every 12 weeks using RECIST and immune-related RECIST (irRECIST) criteria, version 1.1 (PFS-6 and iPFS-6 respectively). Secondary endpoints include assessment of the overall response rate (ORR), clinical benefit rate (CBR), overall survival (OS), and progression free survival (PFS) using RECIST and irRECIST criteria.

PFS was measured from the date of treatment initiation to disease progression or death from any cause, whichever came first. Patients without documented disease progression on their last follow-up were censored at the date of last contact. OS was measured from the date of treatment initiation to the date of death of any cause with patients last known to be alive censored at the date of last contact. PFS and OS were estimated using the Kaplan-Meier method and compared using log-rank tests. All analyses were done using R version 4.0.3.

## Results

### Patient characteristics

A total of twenty-four patients were included: nineteen patients in the ACC cohort and five patients in the other salivary neoplasm cohort. Histologies seen in the salivary neoplasm cohort include parotid gland adenocarcinomas (n=3), salivary duct carcinoma (n=1), and high-grade myoepithelial carcinoma arising from a low-grade biphasic salivary carcinoma (n=1) (Table 1). Among the ACC patients (n=19), the median age was 56.0 years with 47.4% of patients being men. For the salivary gland neoplasm cohort (n=5), the median age was 62.0 years with 60.0% of patients being men. Complete blood counts prior to treatment initiation were collected for 23 patients and molecular characterization performed as part of routine care was available for four patients.

**Table 1:** Summary of patient demographics and characteristics.

|  | **Overall, N = 24** | **Non-ACC, N = 5** | **ACC, N = 19** |
| --- | --- | --- | --- |
| **Mean age (SD)** | 56.3 (14.7) | 60.6 (12.4) | 55.2 (15.3) |
| **Male, n (%)** | 12 (50.0%) | 3 (60.0%) | 9 (47.4%) |
| **BMI (SD)** | 25.4 (3.8) | 27.8 (4.3) | 24.8 (3.5) |
| **ECOG status** |  |  |  |
| **0** | 16 (66.7%) | 4 (80.0%) | 12 (63.2%) |
| **1** | 8 (33.33%) | 1 (20.0%) | 7 (36.8%) |
| **Location** |  |  |  |
| **Salivary gland** | 12 (50.0%) | 4 (80.0%) | 8 (42.1%) |
| **Sinus** | 2 (8.3%) | 0 (0.0%) | 2 (10.5%) |
| **Lacrimal gland** | 3 (12.5%) | 0 (0.0%) | 3 (15.8%) |
| **Base of tongue** | 2 (8.3%) | 0 (0.0%) | 2 (10.5%) |
| **Retromolar trigone** | 1 (4.2%) | 1 (4.2%) | 0 (0.0%) |
| **Nasal cavity** | 1 (4.2%) | 0 (0.0%) | 1 (5.3%) |
| **Mandible** | 1 (4.2%) | 0 (0.0%) | 1 (5.3%) |
| **Ear** | 1 (4.2%) | 0 (0.0%) | 1 (5.3%) |
| **Bartholin gland** | 1 (4.2%) | 0 (0.0%) | 1 (5.3%) |
| **Staging** |  |  |  |
| **Recurrent** | 1 (4.2%) | 0 (0.0%) | 1 (5.3%) |
| **IV** | 23 (95.8%) | 5 100.0%) | 18 (94.7%) |

Abbreviations: SD, standard deviation; N, number

### Outcomes

Among the nineteen ACC patients, the median OS was 30.0 months (95% confidence interval (CI) 15.3 months to not reached), the median PFS was 8.3 months (95% CI 5.5 - 30.0 months), and the disease control rate (DCR) was 42.1% (8/19) using RECIST measurements (Figure 1, Table 2). The PFS-6 and iPFS-6 were both 63% (12/19), whereas OS at 6 months was 89% (17/19). The overall response rate (ORR) in the ACC group was 5% (CR 0%, n=0; confirmed PR 5%, n=1) using both RECIST and irRECIST criteria. Responses of note include a durable PR lasting over 48 months and a patient with ongoing SD lasting over 59 months.

**Table 2:** Best responses.

| **RECIST responses** |  |  |  |
| --- | --- | --- | --- |
| **Characteristic** | **Overall, N = 24** | **Non-ACC, N = 5** | **ACC, N = 19** |
| **CR** | 0 (0.00%) | 0 (0.00%) | 0 (0.00%) |
| **PR** | 1 (4.17%) | 0 (0.00%) | 1 (5.26%) |
| **SD** | 13 (54.17%) | 2 (40.00%) | 11 (57.89%) |
| **PD** | 9 (37.50%) | 3 (60.00%) | 6 (31.58%) |
| **N/A** | 1 (4.17%) | 0 (0.00%) | 1 (5.26%) |
| **DCR\*** | 10 (41.7%) | 2 (40.0%) | 8 (42.1%) |
| **irRECIST responses** |  |  |  |
| **Characteristic** | **Overall, N = 24** | **Non-ACC, N = 5** | **ACC, N = 19** |
| **CR** | 0 (0.00%) | 0 (0.00%) | 0 (0.00%) |
| **PR** | 1 (4.17%) | 0 (0.00%) | 1 (5.26%) |
| **SD** | 15 (62.5%) | 2 (40.00%) | 13 (68.4%) |
| **PD** | 6 (25.0%) | 3 (60.00%) | 3 (15.8%) |
| **N/A** | 2 (8.33%) | 0 (0.00%) | 2 (10.5%) |
| **DCR\*** | 11 (45.8%) | 2 (40.0%) | 9 (47.3%) |

Disease control rate: CR + PR + SD lasting over 24 weeks
Abbreviations: CR, complete response; PR, partial response; SD, stable disease; PD, progressive disease.

**Figure 1.**
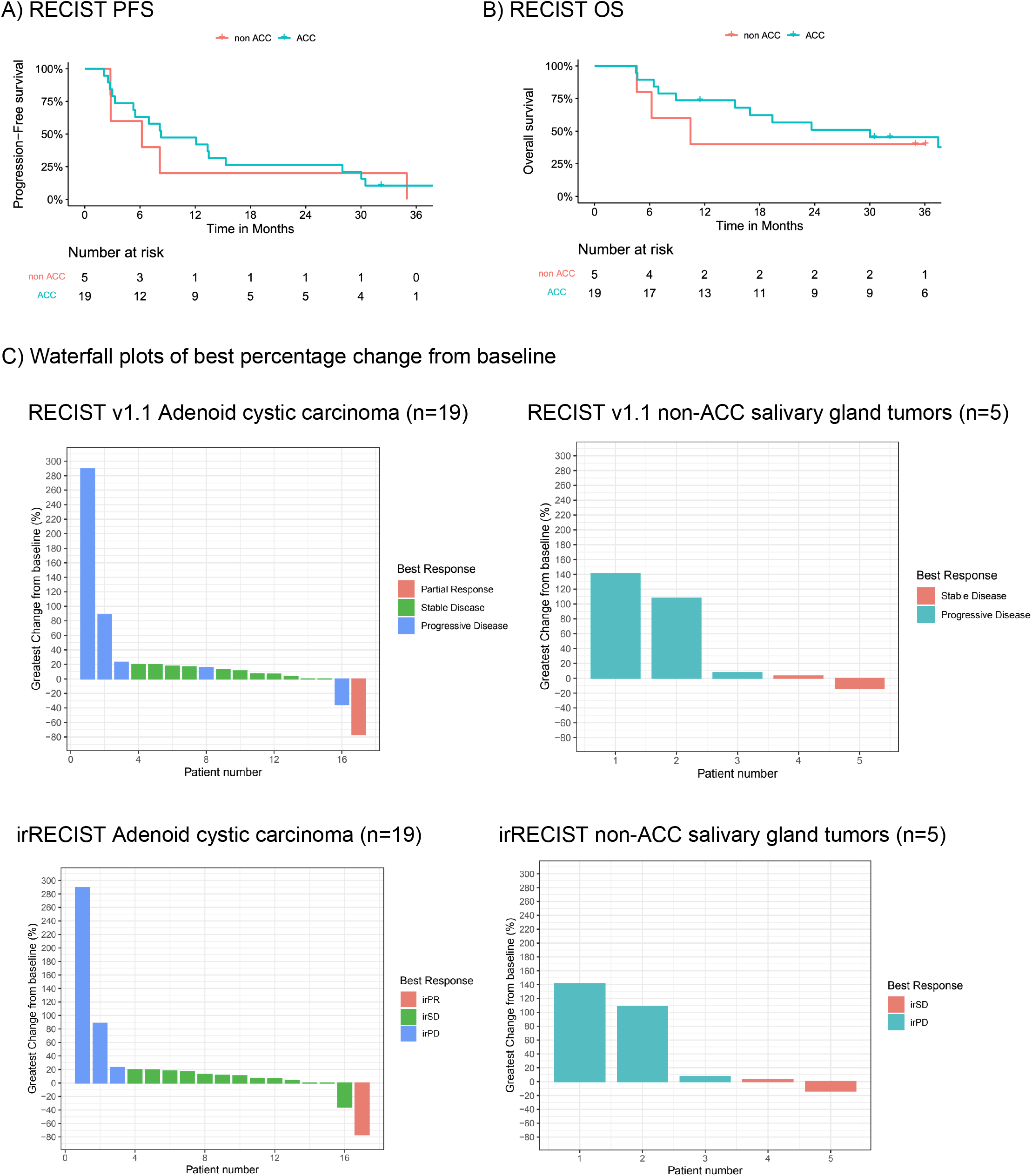
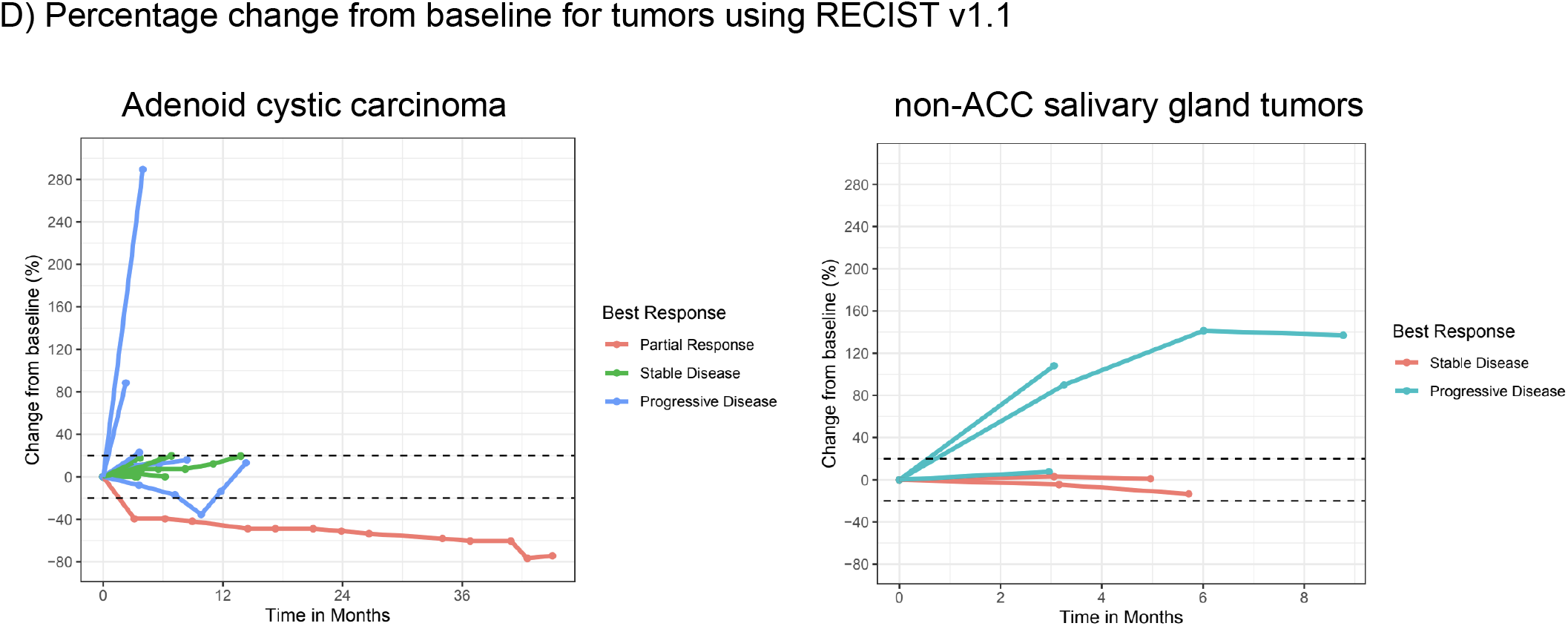
Outcomes of patients with metastatic/recurrent ACC or non-ACC salivary gland cancers treated with nivolumab and ipilimumab. **A)** OS and PFS displayed on Kaplan-Meier curve. **B)** Waterfall plot depicting RECIST or irRECIST progression, stable disease, or partial response. **C)** Spider plot displaying percentage change in longest tumor diameters over time

For the five salivary gland neoplasm patients, the median OS was 10.4 months (95% CI 6.21 months to not reached), the median PFS time was 6.21 months (95% CI 2.83 months to not reached), and the DCR was 40% (2/5). PFS-6 and iPFS-6 were both 40% (2/5) with an OS at 6 months of 80% (4/5) and ORR of 0% using both RECIST and irRECIST criteria (Figure 1, Table 2).

### Toxicities

All 24 of our patients experienced a treatment-related adverse event (AE), the most common of which include fatigue (54%, n=13), lymphocytopenia (46%, n=11), and anemia (42%, n=10). 45.8% of patients (n=11) experienced a grade III-IV treatment-related AE, none of the patients had a grade V treatment-related AE (Table 3, full list of all AEs in supplementary Table 1). A couple of serious treatment-related adverse events were noted including adrenal insufficiency (n=2, one grade II and one grade II), ALT elevation (n=1, grade IV), AST elevation (n=1, grade IV), anorexia (n=1, grade III), and colitis (n=1, grade III). Fortunately, no patient deaths were associated with the study. Lastly, out of all patients who experienced an AE, 12.5% (n=3) led to discontinuation of treatment.

**Table 3:**
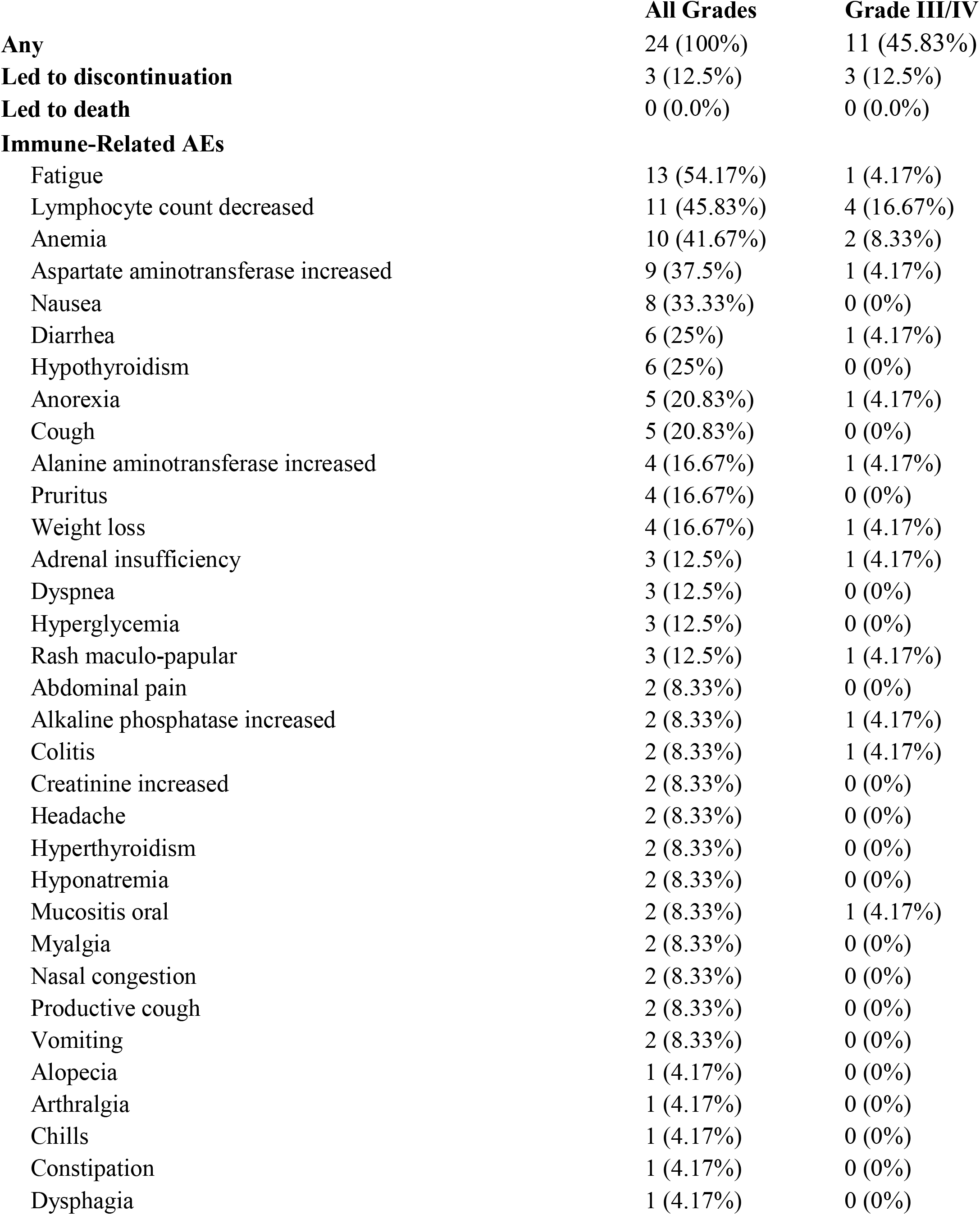
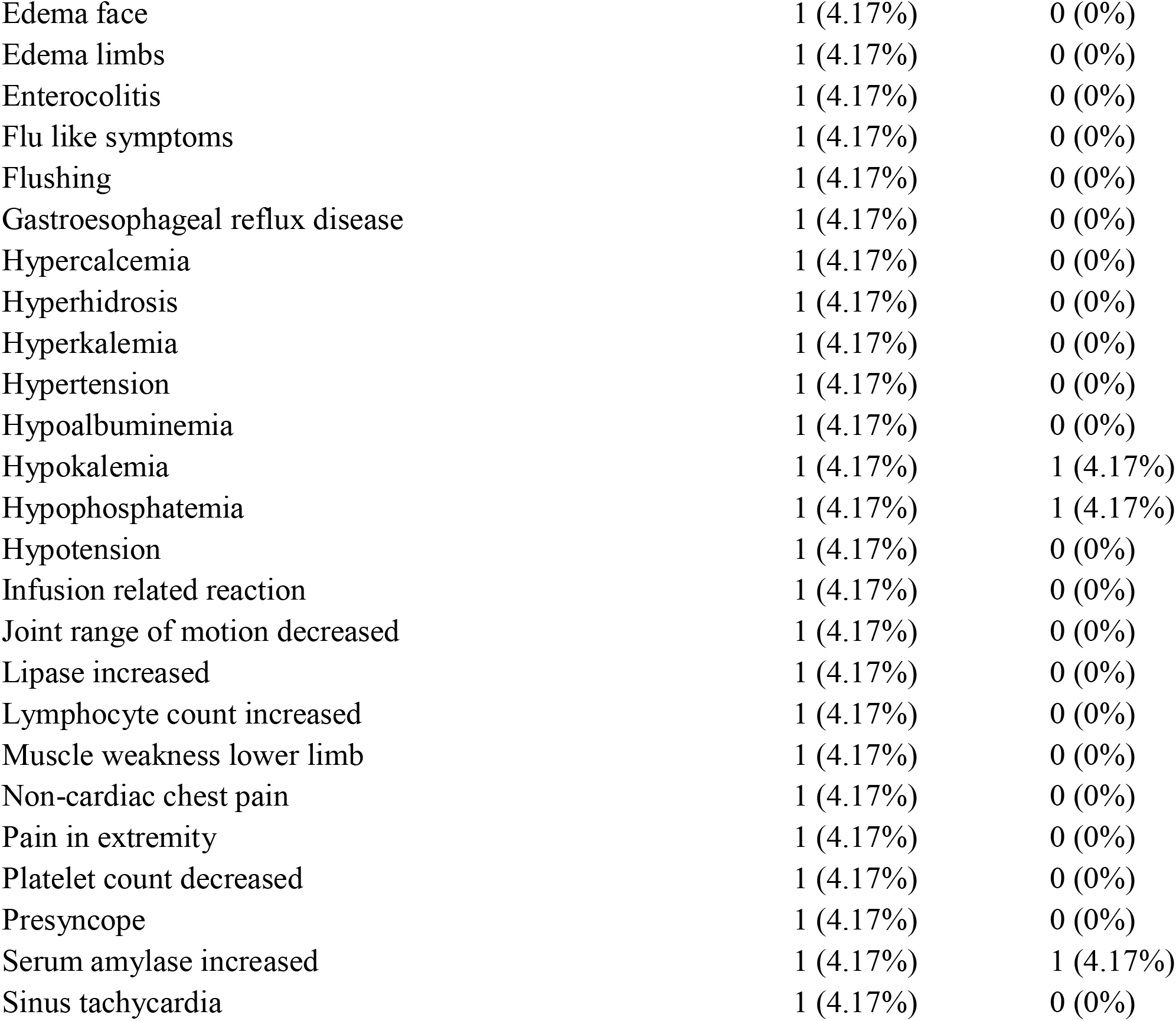
Immune-related adverse events (AEs) separated by grades (I/II vs III/IV/V)

|  | **All Grades** | **Grade III/IV** |
| --- | --- | --- |
| **Any** | 24 (100%) | 11 (45.83%) |
| **Led to discontinuation** | 3 (12.5%) | 3 (12.5%) |
| **Led to death** | 0 (0.0%) | 0 (0.0%) |
| **Immune-Related AEs** |  |  |
| Fatigue | 13 (54.17%) | 1 (4.17%) |
| Lymphocyte count decreased | 11 (45.83%) | 4 (16.67%) |
| Anemia | 10 (41.67%) | 2 (8.33%) |
| Aspartate aminotransferase increased | 9 (37.5%) | 1 (4.17%) |
| Nausea | 8 (33.33%) | 0 (0%) |
| Diarrhea | 6 (25%) | 1 (4.17%) |
| Hypothyroidism | 6 (25%) | 0 (0%) |
| Anorexia | 5 (20.83%) | 1 (4.17%) |
| Cough | 5 (20.83%) | 0 (0%) |
| Alanine aminotransferase increased | 4 (16.67%) | 1 (4.17%) |
| Pruritus | 4 (16.67%) | 0 (0%) |
| Weight loss | 4 (16.67%) | 1 (4.17%) |
| Adrenal insufficiency | 3 (12.5%) | 1 (4.17%) |
| Dyspnea | 3 (12.5%) | 0 (0%) |
| Hyperglycemia | 3 (12.5%) | 0 (0%) |
| Rash maculo-papular | 3 (12.5%) | 1 (4.17%) |
| Abdominal pain | 2 (8.33%) | 0 (0%) |
| Alkaline phosphatase increased | 2 (8.33%) | 1 (4.17%) |
| Colitis | 2 (8.33%) | 1 (4.17%) |
| Creatinine increased | 2 (8.33%) | 0 (0%) |
| Headache | 2 (8.33%) | 0 (0%) |
| Hyperthyroidism | 2 (8.33%) | 0 (0%) |
| Hyponatremia | 2 (8.33%) | 0 (0%) |
| Mucositis oral | 2 (8.33%) | 1 (4.17%) |
| Myalgia | 2 (8.33%) | 0 (0%) |
| Nasal congestion | 2 (8.33%) | 0 (0%) |
| Productive cough | 2 (8.33%) | 0 (0%) |
| Vomiting | 2 (8.33%) | 0 (0%) |
| Alopecia | 1 (4.17%) | 0 (0%) |
| Arthralgia | 1 (4.17%) | 0 (0%) |
| Chills | 1 (4.17%) | 0 (0%) |
| Constipation | 1 (4.17%) | 0 (0%) |
| Dysphagia | 1 (4.17%) | 0 (0%) |
| Edema face | 1 (4.17%) | 0 (0%) |
| Edema limbs | 1 (4.17%) | 0 (0%) |
| Enterocolitis | 1 (4.17%) | 0 (0%) |
| Flu like symptoms | 1 (4.17%) | 0 (0%) |
| Flushing | 1 (4.17%) | 0 (0%) |
| Gastroesophageal reflux disease | 1 (4.17%) | 0 (0%) |
| Hypercalcemia | 1 (4.17%) | 0 (0%) |
| Hyperhidrosis | 1 (4.17%) | 0 (0%) |
| Hyperkalemia | 1 (4.17%) | 0 (0%) |
| Hypertension | 1 (4.17%) | 0 (0%) |
| Hypoalbuminemia | 1 (4.17%) | 0 (0%) |
| Hypokalemia | 1 (4.17%) | 1 (4.17%) |
| Hypophosphatemia | 1 (4.17%) | 1 (4.17%) |
| Hypotension | 1 (4.17%) | 0 (0%) |
| Infusion related reaction | 1 (4.17%) | 0 (0%) |
| Joint range of motion decreased | 1 (4.17%) | 0 (0%) |
| Lipase increased | 1 (4.17%) | 0 (0%) |
| Lymphocyte count increased | 1 (4.17%) | 0 (0%) |
| Muscle weakness lower limb | 1 (4.17%) | 0 (0%) |
| Non-cardiac chest pain | 1 (4.17%) | 0 (0%) |
| Pain in extremity | 1 (4.17%) | 0 (0%) |
| Platelet count decreased | 1 (4.17%) | 0 (0%) |
| Presyncope | 1 (4.17%) | 0 (0%) |
| Serum amylase increased | 1 (4.17%) | 1 (4.17%) |
| Sinus tachycardia | 1 (4.17%) | 0 (0%) |

### Cellular and Molecular characterization

Prior to treatment initiation, twenty-three out of the twenty-four patients had complete blood counts drawn, from which medians of the platelet count (246K/uL), neutrophil count (4.3K/uL), lymphocyte count (1.2K/uL), and neutrophil-to-lymphocyte ratio (3.36) were calculated. Across both ACC and salivary neoplasm cohorts, platelet counts above the median were significantly associated with better OS (p=0.032) and PFS (p=0.046) (Supplemental Figure 1). Additionally, although not significant, there was a slight trend for improved OS and PFS among patients with neutrophil-to-lymphocyte ratios greater than five (OS p=0.42, PFS p=0.25) and above median neutrophil counts (OS p=0.24, PFS p=0.18).

Four of the twenty-four patients additionally underwent next-generation sequencing (NGS), all of whom had at least two pathological aberrations detected. Of special note, two MYB-NFIB translocations, one NOTCH1 Cys1383 frameshift mutation, one MTOR N1760K mutation, one FGFR2 overexpression, one HRAS overexpression, and one PDGFRA copy number gain were detected (Supplementary Table 2). None of the patients had high levels of tumor mutational burden detected. Additionally, the two patients who also had available PDL1 immunohistochemistry available had <1% staining.

Six of the twenty-four patients had H&E-stained whole slide images (WSIs) available for analysis using an artificial intelligence (AI)-powered WSI analyzer, Lunit SCOPE IO (Lunit, Seoul, Republic of Korea), which analyzes the spatial distribution of tumor infiltrating lymphocytes (TILs) throughout the WSI to predict responsiveness to ICI[15,16]. Among these six patients, the three who reached SD had the lowest inflamed scores and were categorized as immune desert, corresponding to below-threshold concentrations of TILs throughout the tumor microenvironment (Table 4). In contrast, the remaining three patients, who had PD as the best response, showed diverse phenotype: two were categorized as immune-excluded, representing higher TIL concentration in the cancer stroma area, and one case was classified as immune-desert, despite having relatively higher TIL concentration in both the cancer and cancer stroma areas.

**Table 4:**
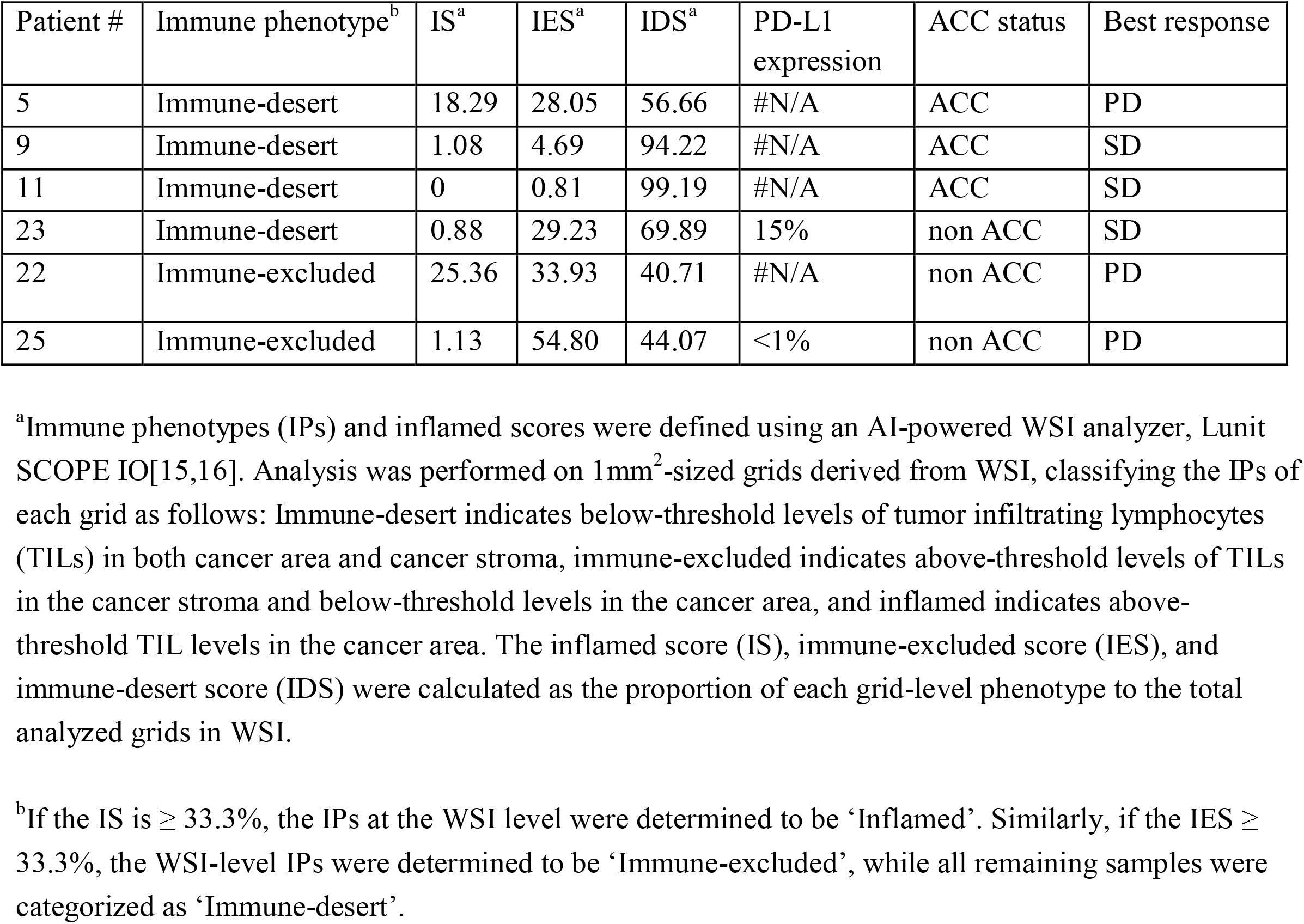
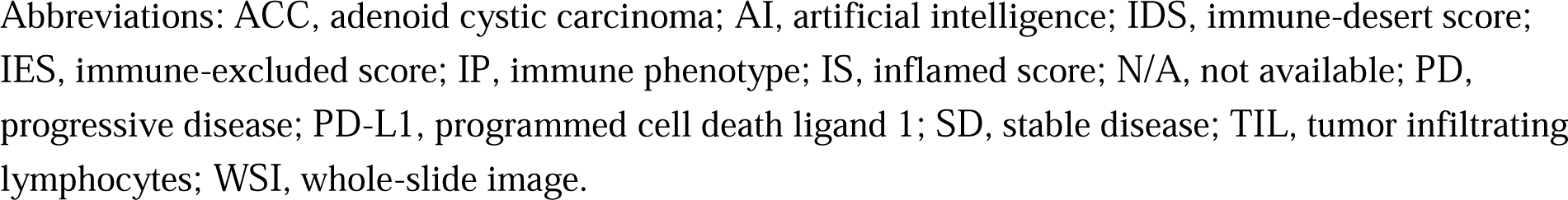
Immune phenotype of patients with available H&E slides.

| Patient # | Immune phenotype<sup>b</sup> | IS<sup>a</sup> | IES<sup>a</sup> | IDS<sup>a</sup> | PD-L1 expression | ACC status | Best response |
| --- | --- | --- | --- | --- | --- | --- | --- |
| 5 | Immune-desert | 18.29 | 28.05 | 56.66 | #N/A | ACC | PD |
| 9 | Immune-desert | 1.08 | 4.69 | 94.22 | #N/A | ACC | SD |
| 11 | Immune-desert | 0 | 0.81 | 99.19 | #N/A | ACC | SD |
| 23 | Immune-desert | 0.88 | 29.23 | 69.89 | 15% | non ACC | SD |
| 22 | Immune-excluded | 25.36 | 33.93 | 40.71 | #N/A | non ACC | PD |
| 25 | Immune-excluded | 1.13 | 54.80 | 44.07 | <1% | non ACC | PD |

<sup>a</sup>Immune phenotypes (IPs) and inflamed scores were defined using an AI-powered WSI analyzer, Lunit SCOPE IO[15,16]. Analysis was performed on 1mm<sup>2</sup>-sized grids derived from WSI, classifying the IPs of each grid as follows: Immune-desert indicates below-threshold levels of tumor infiltrating lymphocytes (TILs) in both cancer area and cancer stroma, immune-excluded indicates above-threshold levels of TILs in the cancer stroma and below-threshold levels in the cancer area, and inflamed indicates above-threshold TIL levels in the cancer area. The inflamed score (IS), immune-excluded score (IES), and immune-desert score (IDS) were calculated as the proportion of each grid-level phenotype to the total analyzed grids in WSI.
<sup>b</sup>If the IS is $\geq 33.3\%$ , the IPs at the WSI level were determined to be ‘Inflamed’. Similarly, if the IES $\geq 33.3\%$ , the WSI-level IPs were determined to be ‘Immune-excluded’, while all remaining samples were categorized as ‘Immune-desert’.
Abbreviations: ACC, adenoid cystic carcinoma; AI, artificial intelligence; IDS, immune-desert score; IES, immune-excluded score; IP, immune phenotype; IS, inflamed score; N/A, not available; PD, progressive disease; PD-L1, programmed cell death ligand 1; SD, stable disease; TIL, tumor infiltrating lymphocytes; WSI, whole-slide image.

## Discussion

In this study, nivolumab and ipilimumab had a moderate efficacy for patients with recurrent/metastatic ACC, while patients with other salivary gland tumors did not experience responses to the combined checkpoint inhibition. This trial’s results complement two previous studies that evaluated nivolumab/ipilimumab as treatment for patients with recurrent/metastatic ACC or other salivary gland cancers. Compared to the 5% (1/19) ORR in our study’s ACC cohort, our previous study and others found ORRs of 6% (2/32) and 4% (1/26) with comparably durable responses (median PFS 4.4 months vs 8.3 months in this study) in ACC patients [13,14]. In our study’s non-ACC salivary carcinoma cohort, we saw a 0% (0/5) ORR, whereas prior trials saw 16% (5/32) and 9% (3/35) ORRs in patients with recurrent/metastatic salivary gland cancers also with comparably durable responses (median PFS 4.6 months vs. 6.21 months in our study) [14,17]. The few durable responses seen in our current and prior trials were also seen in a case report in which a patient with recurrent and metastatic parotid gland cystadenocarcinoma had a complete response to nivolumab and ipilimumab[18]. These studies highlight how the majority of patients with ACC and other salivary gland cancer do not respond well to immune checkpoint inhibition. However, a unique subgroup of patients do respond well, and understanding the reasons underlying their responses is the next step.

Prior studies have also evaluated PD-1/PD-L1 inhibitor monotherapy for patients with both ACC and non-ACC salivary gland cancers. Among the ACC-related studies, a trial comparing pembrolizumab with and without radiation for patients with recurrent/metastatic ACC saw an ORR of 0% for all patients included[19]. A separate trial evaluating nivolumab included 46 patients with recurrent/metastatic ACC saw an 8.7% ORR[12]. These studies yielded comparable ORRs to our study and other prior dual checkpoint inhibitor trials for recurrent/metastatic ACC. However, their median PFS (4.9 and 6.6 for pembrolizumab only) were lower than that seen in our study (8.3 months), suggesting a possible role of CTLA-4 inhibitors like ipilimumab in extending the responses that patients have to immunotherapy[20,21].

Non-ACC salivary gland carcinomas were also investigated in past trials, including nivolumab monotherapy for patients with recurrent or metastatic salivary gland carcinomas that resulted in ORRs ranging from 4.2 to 8.7%[12,22,23]. Another subset of trials evaluated pembrolizumab monotherapy, achieving ORRs of 12% and 4.6% in patients with recurrent/metastatic salivary gland carcinomas[24,25]. Although PD-1/PD-L1 inhibitor monotherapy yielded comparable ORRs to our study, their reported median PFS were also lower (1.6 - 4.9 months) compared to our study (6.1 months)[12,22–25]. This again suggests a role of ipilimumab in prolonging responses to PD-1/PD-L1 therapy, although durable positive responses over 48 weeks were seen in some monotherapy studies[18,21,25]

In addition to data on overall responses, we explored molecular landscapes to identify potential markers for treatment outcomes in the four patients with sequencing performed. Our one patient with ACC who displayed a positive response had NGS-based testing done, which identified two frameshift mutations in genes, BAP1 and BCOR. These genes are associated with inflammatory tumor microenvironments and improved immune checkpoint inhibitor responses respectively when mutated in different types of lung cancer, and therefore could explain the exceptional benefit the patient saw with nivolumab/ipilimumab treatment [26,27].

Another set of markers that we found to have correlation with immune checkpoint inhibitor outcomes were pre-treatment cell counts for platelets, neutrophils, and lymphocytes. Platelet counts especially were notable for their association with treatment outcomes: patients at below-median platelet counts had a significant association with increased PFS and OS (p=0.032 and 0.046 respectively) with hazard ratio 3.60 (95% CI 1.11-11.7) for increased OS. The protective association with low platelets has been previously described, as platelets have been known to promote tumor progression through angiogenesis and adhesion molecule production[28]. Furthermore, the association between elevated platelet counts and neutrophil/lymphocyte ratios with poor outcomes has already been described for many other cancers[29]. However, their replicability in our study tells us that the host immune milieu is important no matter the tumor type, including for ACC and other salivary gland cancers.

We also explored a subset of our patients with available tissue slides for TIL distribution and found that all three patients with lower TIL concentrations reached SD, whereas the other three patients only had PD while on ICI treatment. Previous literature describes how immune checkpoint blockade is correlated with an increase in intratumoral lymphocyte concentration[30]. In the case of CTLA-4 inhibition, pre-treatment tumor T-lymphocyte levels were not found to correlate with outcomes in patients with melanoma[30]. Instead, clinical activity was correlated with the increase in T lymphocyte levels after treatment. In our study, patients only had pre-treatment samples analyzed. Thus, a possible explanation for patients having more durable responses with lower pre-ICI treatment lymphocyte levels is that they benefited from T lymphocyte recruitment after combined PD-1/CTLA-4 inhibition. Nonetheless, there is a need for further investigation using a larger cohort size.

Our study is an innovative basket trial whose main strengths include the NGS and histopathologic data available for our unique positive responder and a handful of other patients. This lends a better understanding of the few patients who do respond to immune checkpoint therapy. One main limitation in working with patients who have ACC and salivary gland tumors is that their cancer biology is heterogeneous and contain many distinct histologic subtypes that are not controlled for, making it difficult to draw similarities between studies. Another weakness is the lack of a control group. Due to the naturally indolent nature of ACC growth and the tendency for delayed recurrence after treatment, a control group would help put this trial’s patient responses into perspective of the specific clinical courses commonly seen in ACC patients[6]. Additional limitations include small sample sizes, limited molecular characterization of patient samples, and the lack of patient randomization.

In conclusion, ACC and other salivary gland cancers compose a group of rare heterogenous malignancies that have no currently FDA-approved treatment options. Treatment with nivolumab and ipilimumab results in modest response rates for patients with ACC only with results comparable to previously conducted studies on dual checkpoint inhibition[13,17] and PD-1/PD-L1 monotherapy[12,19,22–25]. We investigated further into a subset of our patients to examine their genomic, transcriptomic, and hematologic profiles. The one patient that had a PR was found to express two mutations that have established associations with the immune microenvironment, suggesting a possible cause for their elevated response. Further studies are warranted to investigate ACC and other salivary gland cancer biology and how well it predicts immune checkpoint therapy outcomes.

## Data availability statement

All data pertinent to our study has been included in this manuscript or uploaded alongside it as supplementary information.

## Ethics statements

Informed consent was obtained directly from patients for participation in this study and its publication. Study approval was given by the institutional IRB in accordance with federally mandated regulations. Patient informed consent was individually obtained in written form, and patients were not involved in the design, conduct, reporting, or dissemination of this study.

## Acknowledgments

We thank the patients and their families for their participation in this study.

## Author contribution statement

YKC: conceptualization, methodology, investigation, writing, review. RD: data curation and analysis, writing, review. LIC: writing and review. YO and IH: data analysis. BA, SS, SK: data curation and review. DM, VF, MM: conceptualization, methodology, investigation, review

## Works Cited

1 Coca-Pelaz A, Rodrigo JP, Bradley PJ, et al. Adenoid cystic carcinoma of the head and neck--An update. Oral Oncol. 2015;51:652–61.

2 Dillon PM, Petroni GR, Horton BJ, et al. A Phase II Study of Dovitinib in Patients with Recurrent or Metastatic Adenoid Cystic Carcinoma. Clin Cancer Res Off J Am Assoc Cancer Res. 2017;23:4138–45.

3 Chen Y, Zheng Z-Q, Chen F-P, et al. Role of Postoperative Radiotherapy in Nonmetastatic Head and Neck Adenoid Cystic Carcinoma. J Natl Compr Cancer Netw JNCCN. 2020;18:1476–84.

4 Jang S, Patel PN, Kimple RJ, et al. Clinical Outcomes and Prognostic Factors of Adenoid Cystic Carcinoma of the Head and Neck. Anticancer Res. 2017;37:3045–52.

5 Hitre E, Budai B, Takácsi-Nagy Z, et al. Cetuximab and platinum-based chemoradioor chemotherapy of patients with epidermal growth factor receptor expressing adenoid cystic carcinoma: a phase II trial. Br J Cancer. 2013;109:1117–22.

6 Chae YK, Chung SY, Davis AA, et al. Adenoid cystic carcinoma: current therapy and potential therapeutic advances based on genomic profiling. Oncotarget. 2015;6:37117–34.

7 Miller LE, Au V, Mokhtari TE, et al. A Contemporary Review of Molecular Therapeutic Targets for Adenoid Cystic Carcinoma. Cancers. 2022;14:992.

8 Meng Y, Jin J, Gong C, et al. Phase II study of chidamide in combination with cisplatin in patients with metastatic triple-negative breast cancer. Ann Palliat Med. 2021;10:11255–64.

9 Ahn, Myung-Ju, Kang, Hyunseok, Muzaffar, Jameel, et al. A phase 2 study of the oral vascular endothelial growth factor receptor 2 (VEGFR2) inhibitor, rivoceranib, for recurrent or metastatic (R/M) adenoid cystic carcinoma (ACC). J Clin Oncol. 2022;40:6020–6020.

10 Goncalves PH, Heilbrun LK, Barrett MT, et al. A phase 2 study of vorinostat in locally advanced, recurrent, or metastatic adenoid cystic carcinoma. Oncotarget. 2017;8:32918–29.

11 Kooshkaki O, Derakhshani A, Hosseinkhani N, et al. Combination of Ipilimumab and Nivolumab in Cancers: From Clinical Practice to Ongoing Clinical Trials. Int J Mol Sci. 2020;21:4427.

12 Fayette J, Even C, Digue L, et al. NISCAHN: A phase II, multicenter nonrandomized trial aiming at evaluating nivolumab (N) in two cohorts of patients (pts) with recurrent/metastatic (R/M) salivary gland carcinoma of the head and neck (SGCHN), on behalf of the Unicancer Head & Neck Group. J Clin Oncol. 2019;37:6083–6083.

13 Tchekmedyian V, Sherman EJ, Dunn L, et al. A phase II trial cohort of nivolumab plus ipilimumab in patients (Pts) with recurrent/metastatic adenoid cystic carcinoma (R/M ACC). J Clin Oncol. 2019;37:6084–6084.

14 Chae YK, Othus M, Patel SP, et al. Abstract 3418: A phase II basket trial of dual anti-CTLA-4 and anti-PD-1 blockade in rare tumors (DART) SWOG S1609: The salivary gland tumor cohort. Cancer Res. 2020;80:3418–3418.

15 Jung HA, Park K-U, Cho S, et al. A Phase II Study of Nivolumab plus Gemcitabine in Patients with Recurrent or Metastatic Nasopharyngeal Carcinoma (KCSG HN17–11). Clin Cancer Res. 2022;28:4240–7.

16 Park S, Ock C-Y, Kim H, et al. Artificial Intelligence–Powered Spatial Analysis of Tumor-Infiltrating Lymphocytes as Complementary Biomarker for Immune Checkpoint Inhibition in Non– Small-Cell Lung Cancer. J Clin Oncol. 2022;40:1916–28.

17 Burman B, Sherman EJ, Dunn L, et al. A phase II trial cohort of nivolumab plus ipilimumab in patients (Pts) with recurrent/metastatic salivary gland cancers (R/M SGCs). J Clin Oncol. 2021;39:6002–6002.

18 Nakamura Y, Nakayama M, Nishimura B, et al. Case Report: Complete Response of Recurrent and Metastatic Cystadenocarcinoma of the Parotid Gland With a Single Course of Combined Nivolumab and Ipilimumab Therapy. Front Oncol. 2021;11:618201.

19 Mahmood U, Bang A, Chen Y-H, et al. A Randomized Phase 2 Study of Pembrolizumab With or Without Radiation in Patients With Recurrent or Metastatic Adenoid Cystic Carcinoma. Int J Radiat Oncol. 2021;109:134–44.

20 Hellmann MD, Paz-Ares L, Bernabe Caro R, et al. Nivolumab plus Ipilimumab in Advanced Non– Small-Cell Lung Cancer. N Engl J Med. 2019;381:2020–31.

21 Johnson ML, Cho BC, Luft A, et al. Durvalumab With or Without Tremelimumab in Combination With Chemotherapy as First-Line Therapy for Metastatic Non–Small-Cell Lung Cancer: The Phase III POSEIDON Study. J Clin Oncol. 2023;41:1213–27.

22 Niwa K, Kawakita D, Nagao T, et al. Multicentre, retrospective study of the efficacy and safety of nivolumab for recurrent and metastatic salivary gland carcinoma. Sci Rep. 2020;10:16988.

23 Nagatani Y, Kiyota N, Yamazaki T, et al. A phase II trial of nivolumab for patients with platinum-refractory recurrent or metastatic salivary gland cancer. J Clin Oncol. 2023;41:6092–6092.

24 Cohen RB, Delord J-P, Doi T, et al. Pembrolizumab for the Treatment of Advanced Salivary Gland Carcinoma: Findings of the Phase 1b KEYNOTE-028 Study. Am J Clin Oncol. 2018;41:1083–8.

25 Even C, Delord J-P, Price KA, et al. Evaluation of pembrolizumab monotherapy in patients with previously treated advanced salivary gland carcinoma in the phase 2 KEYNOTE-158 study. Eur J Cancer. 2022;171:259–68.

26 Ladanyi M, Sanchez Vega F, Zauderer M. Loss of BAP1 as a candidate predictive biomarker for immunotherapy of mesothelioma. Genome Med. 2019;11:18.

27 Liu D, Benzaquen J, Morris LGT, et al. Mutations in KMT2C, BCOR and KDM5C Predict Response to Immune Checkpoint Blockade Therapy in Non-Small Cell Lung Cancer. Cancers. 2022;14:2816.

28 Wojtukiewicz MZ, Sierko E, Hempel D, et al. Platelets and cancer angiogenesis nexus. Cancer Metastasis Rev. 2017;36:249–62.

29 Li B, Zhou P, Liu Y, et al. Platelet-to-lymphocyte ratio in advanced Cancer: Review and metaanalysis. Clin Chim Acta. 2018;483:48–56.

30 Plesca I, Tunger A, Müller L, et al. Characteristics of Tumor-Infiltrating Lymphocytes Prior to and During Immune Checkpoint Inhibitor Therapy. Front Immunol. 2020;11:364.

