## Supplemental methods for "Phase II study of nivolumab and ipilimumab for treatment of metastatic/recurrent adenoid cystic carcinoma (ACC) of all anatomic sites of origin and other malignant salivary gland tumors"

For patients with metastatic/recurrent ACC, the optimal two-stage design to test the null hypothesis that P≤0.450 versus the alternative that P≥0.650 has an expected sample size of 24.70 and a probability of early termination of 0.654. If the drug is not actually effective, there is a 0.048 (target 0.05) probability of concluding that it is effective. If the drug is actually effective, there is a 0.196 (target 0.200) probability of concluding that it is not.
