## Supplemental Table 1 for "Phase II study of nivolumab and ipilimumab for treatment of metastatic/recurrent adenoid cystic carcinoma (ACC) of all anatomic sites of origin and other malignant salivary gland tumors"

Supplemental Table 1: All adverse events (AEs) separated by grades (I/II vs III/IV/V)

|  | **AE** | **All Grades** | **Grade I/II** | **Grade III/IV/V** |
| --- | --- | --- | --- | --- |
| 1 | Hypertension | 24 (100%) | 15 (62.5%) | 9 (37.5%) |
| 2 | Fatigue | 18 (75%) | 17 (70.83%) | 1 (4.17%) |
| 3 | Lymphocyte count decreased | 18 (75%) | 12 (50%) | 6 (25%) |
| 4 | Anemia | 17 (70.83%) | 14 (58.33%) | 3 (12.5%) |
| 5 | Aspartate aminotransferase increased | 15 (62.5%) | 13 (54.17%) | 2 (8.33%) |
| 6 | Alkaline phosphatase increased | 12 (50%) | 9 (37.5%) | 3 (12.5%) |
| 7 | Hypoalbuminemia | 12 (50%) | 12 (50%) | 0 (0%) |
| 8 | Nausea | 12 (50%) | 11 (45.83%) | 1 (4.17%) |
| 9 | Weight loss | 12 (50%) | 10 (41.67%) | 2 (8.33%) |
| 10 | Anorexia | 11 (45.83%) | 8 (33.33%) | 3 (12.5%) |
| 11 | Constipation | 11 (45.83%) | 11 (45.83%) | 0 (0%) |
| 12 | Dyspnea | 10 (41.67%) | 9 (37.5%) | 1 (4.17%) |
| 13 | Cough | 9 (37.5%) | 9 (37.5%) | 0 (0%) |
| 14 | Hyponatremia | 9 (37.5%) | 7 (29.17%) | 2 (8.33%) |
| 15 | Hypothyroidism | 9 (37.5%) | 9 (37.5%) | 0 (0%) |
| 16 | Alanine aminotransferase increased | 8 (33.33%) | 6 (25%) | 2 (8.33%) |
| 17 | Hyperglycemia | 8 (33.33%) | 8 (33.33%) | 0 (0%) |
| 18 | Pain | 8 (33.33%) | 8 (33.33%) | 0 (0%) |
| 19 | Diarrhea | 7 (29.17%) | 6 (25%) | 1 (4.17%) |
| 20 | Gastroesophageal reflux disease | 7 (29.17%) | 7 (29.17%) | 0 (0%) |
| 21 | Trismus | 7 (29.17%) | 7 (29.17%) | 0 (0%) |
| 22 | Abdominal pain | 6 (25%) | 4 (16.67%) | 2 (8.33%) |
| 23 | Back pain | 6 (25%) | 6 (25%) | 0 (0%) |
| 24 | Blood bilirubin increased | 6 (25%) | 5 (20.83%) | 1 (4.17%) |
| 25 | Dry mouth | 6 (25%) | 6 (25%) | 0 (0%) |
| 26 | Hyperthyroidism | 6 (25%) | 6 (25%) | 0 (0%) |
| 27 | Hypokalemia | 6 (25%) | 3 (12.5%) | 3 (12.5%) |
| 28 | Pruritus | 6 (25%) | 6 (25%) | 0 (0%) |
| 29 | Anxiety | 5 (20.83%) | 4 (16.67%) | 1 (4.17%) |
| 30 | Headache | 5 (20.83%) | 4 (16.67%) | 1 (4.17%) |
| 31 | Hypocalcemia | 5 (20.83%) | 5 (20.83%) | 0 (0%) |
| 32 | Rash maculo-papular | 5 (20.83%) | 3 (12.5%) | 2 (8.33%) |
| 33 | Creatinine increased | 4 (16.67%) | 3 (12.5%) | 1 (4.17%) |
| 34 | Insomnia | 4 (16.67%) | 4 (16.67%) | 0 (0%) |
| 35 | Pain in extremity | 4 (16.67%) | 3 (12.5%) | 1 (4.17%) |
| 36 | Paresthesia | 4 (16.67%) | 4 (16.67%) | 0 (0%) |
| 37 | Platelet count decreased | 4 (16.67%) | 4 (16.67%) | 0 (0%) |
| 38 | Vomiting | 4 (16.67%) | 4 (16.67%) | 0 (0%) |
| 39 | Adrenal insufficiency | 3 (12.5%) | 2 (8.33%) | 1 (4.17%) |
| 40 | Colitis | 3 (12.5%) | 1 (4.17%) | 2 (8.33%) |
| 41 | Dysgeusia | 3 (12.5%) | 3 (12.5%) | 0 (0%) |
| 42 | Facial muscle weakness | 3 (12.5%) | 3 (12.5%) | 0 (0%) |
| 43 | Gait disturbance | 3 (12.5%) | 3 (12.5%) | 0 (0%) |
| 44 | Hearing impaired | 3 (12.5%) | 3 (12.5%) | 0 (0%) |
| 45 | Hypoglycemia | 3 (12.5%) | 3 (12.5%) | 0 (0%) |
| 46 | Mucositis oral | 3 (12.5%) | 2 (8.33%) | 1 (4.17%) |
| 47 | Myalgia | 3 (12.5%) | 3 (12.5%) | 0 (0%) |
| 48 | Nasal congestion | 3 (12.5%) | 3 (12.5%) | 0 (0%) |
| 49 | Non-cardiac chest pain | 3 (12.5%) | 2 (8.33%) | 1 (4.17%) |
| 50 | Productive cough | 3 (12.5%) | 3 (12.5%) | 0 (0%) |
| 51 | Sinus tachycardia | 3 (12.5%) | 3 (12.5%) | 0 (0%) |
| 52 | Urinary frequency | 3 (12.5%) | 3 (12.5%) | 0 (0%) |
| 53 | Urinary tract infection | 3 (12.5%) | 3 (12.5%) | 0 (0%) |
| 54 | Weight gain | 3 (12.5%) | 3 (12.5%) | 0 (0%) |
| 55 | Amnesia | 2 (8.33%) | 2 (8.33%) | 0 (0%) |
| 56 | Bone pain | 2 (8.33%) | 2 (8.33%) | 0 (0%) |
| 57 | Confusion | 2 (8.33%) | 1 (4.17%) | 1 (4.17%) |
| 58 | Depression | 2 (8.33%) | 2 (8.33%) | 0 (0%) |
| 59 | Dermatitis radiation | 2 (8.33%) | 2 (8.33%) | 0 (0%) |
| 60 | Ear pain | 2 (8.33%) | 2 (8.33%) | 0 (0%) |
| 61 | Edema limbs | 2 (8.33%) | 2 (8.33%) | 0 (0%) |
| 62 | Fever | 2 (8.33%) | 2 (8.33%) | 0 (0%) |
| 63 | Flu like symptoms | 2 (8.33%) | 2 (8.33%) | 0 (0%) |
| 64 | Hyperkalemia | 2 (8.33%) | 2 (8.33%) | 0 (0%) |
| 65 | Hypotension | 2 (8.33%) | 2 (8.33%) | 0 (0%) |
| 66 | INR increased | 2 (8.33%) | 2 (8.33%) | 0 (0%) |
| 67 | Lipase increased | 2 (8.33%) | 2 (8.33%) | 0 (0%) |
| 68 | Neck pain | 2 (8.33%) | 1 (4.17%) | 1 (4.17%) |
| 69 | Peripheral sensory neuropathy | 2 (8.33%) | 2 (8.33%) | 0 (0%) |
| 70 | Pleural effusion | 2 (8.33%) | 1 (4.17%) | 1 (4.17%) |
| 71 | Presyncope | 2 (8.33%) | 2 (8.33%) | 0 (0%) |
| 72 | Sepsis | 2 (8.33%) | 0 (0%) | 2 (8.33%) |
| 73 | Sinusitis | 2 (8.33%) | 1 (4.17%) | 1 (4.17%) |
| 74 | Superficial soft tissue fibrosis | 2 (8.33%) | 2 (8.33%) | 0 (0%) |
| 75 | Thromboembolic event | 2 (8.33%) | 0 (0%) | 2 (8.33%) |
| 76 | Tumor pain | 2 (8.33%) | 1 (4.17%) | 1 (4.17%) |
| 77 | Acute kidney injury | 1 (4.17%) | 0 (0%) | 1 (4.17%) |
| 78 | Alkalosis | 1 (4.17%) | 0 (0%) | 1 (4.17%) |
| 79 | Allergic rhinitis | 1 (4.17%) | 1 (4.17%) | 0 (0%) |
| 80 | Alopecia | 1 (4.17%) | 1 (4.17%) | 0 (0%) |
| 81 | Arthralgia | 1 (4.17%) | 1 (4.17%) | 0 (0%) |
| 82 | Arthritis | 1 (4.17%) | 1 (4.17%) | 0 (0%) |
| 83 | Chest wall pain | 1 (4.17%) | 0 (0%) | 1 (4.17%) |
| 84 | Chills | 1 (4.17%) | 1 (4.17%) | 0 (0%) |
| 85 | Cholesterol high | 1 (4.17%) | 1 (4.17%) | 0 (0%) |
| 86 | Cognitive disturbance | 1 (4.17%) | 0 (0%) | 1 (4.17%) |
| 87 | Dehydration | 1 (4.17%) | 1 (4.17%) | 0 (0%) |
| 88 | Dental caries | 1 (4.17%) | 1 (4.17%) | 0 (0%) |
| 89 | Dyspepsia | 1 (4.17%) | 1 (4.17%) | 0 (0%) |
| 90 | Dysphagia | 1 (4.17%) | 1 (4.17%) | 0 (0%) |
| 91 | Edema face | 1 (4.17%) | 1 (4.17%) | 0 (0%) |
| 92 | Edema trunk | 1 (4.17%) | 1 (4.17%) | 0 (0%) |
| 93 | Enterocolitis | 1 (4.17%) | 1 (4.17%) | 0 (0%) |
| 94 | Erectile dysfunction | 1 (4.17%) | 1 (4.17%) | 0 (0%) |
| 95 | Erythema multiforme | 1 (4.17%) | 1 (4.17%) | 0 (0%) |
| 96 | Eyelid function disorder | 1 (4.17%) | 1 (4.17%) | 0 (0%) |
| 97 | Fall | 1 (4.17%) | 1 (4.17%) | 0 (0%) |
| 98 | Fibrinogen decreased | 1 (4.17%) | 1 (4.17%) | 0 (0%) |
| 99 | Flank pain | 1 (4.17%) | 1 (4.17%) | 0 (0%) |
| 100 | Flushing | 1 (4.17%) | 1 (4.17%) | 0 (0%) |
| 101 | Fracture | 1 (4.17%) | 1 (4.17%) | 0 (0%) |
| 102 | Gastroparesis | 1 (4.17%) | 1 (4.17%) | 0 (0%) |
| 103 | Hematuria | 1 (4.17%) | 1 (4.17%) | 0 (0%) |
| 104 | Hiccups | 1 (4.17%) | 1 (4.17%) | 0 (0%) |
| 105 | Hypercalcemia | 1 (4.17%) | 1 (4.17%) | 0 (0%) |
| 106 | Hyperhidrosis | 1 (4.17%) | 1 (4.17%) | 0 (0%) |
| 107 | Hypernatremia | 1 (4.17%) | 1 (4.17%) | 0 (0%) |
| 108 | Hypophosphatemia | 1 (4.17%) | 0 (0%) | 1 (4.17%) |
| 109 | Infusion related reaction | 1 (4.17%) | 1 (4.17%) | 0 (0%) |
| 110 | Joint range of motion decreased | 1 (4.17%) | 1 (4.17%) | 0 (0%) |
| 111 | Laryngitis | 1 (4.17%) | 1 (4.17%) | 0 (0%) |
| 112 | Lip pain | 1 (4.17%) | 1 (4.17%) | 0 (0%) |
| 113 | Lymphocyte count increased | 1 (4.17%) | 1 (4.17%) | 0 (0%) |
| 114 | Muscle weakness lower limb | 1 (4.17%) | 1 (4.17%) | 0 (0%) |
| 115 | Muscle weakness right-sided | 1 (4.17%) | 0 (0%) | 1 (4.17%) |
| 116 | Osteonecrosis of jaw | 1 (4.17%) | 1 (4.17%) | 0 (0%) |
| 117 | Periodontal disease | 1 (4.17%) | 0 (0%) | 1 (4.17%) |
| 118 | Portal vein thrombosis | 1 (4.17%) | 1 (4.17%) | 0 (0%) |
| 119 | Postnasal drip | 1 (4.17%) | 1 (4.17%) | 0 (0%) |
| 120 | Rash acneiform | 1 (4.17%) | 1 (4.17%) | 0 (0%) |
| 121 | Respiratory failure | 1 (4.17%) | 0 (0%) | 1 (4.17%) |
| 122 | Seizure | 1 (4.17%) | 0 (0%) | 1 (4.17%) |
| 123 | Serum amylase increased | 1 (4.17%) | 0 (0%) | 1 (4.17%) |
| 124 | Skin hyperpigmentation | 1 (4.17%) | 1 (4.17%) | 0 (0%) |
| 125 | Skin infection | 1 (4.17%) | 1 (4.17%) | 0 (0%) |
| 126 | Somnolence | 1 (4.17%) | 1 (4.17%) | 0 (0%) |
| 127 | Sore throat | 1 (4.17%) | 1 (4.17%) | 0 (0%) |
| 128 | Spinal fracture | 1 (4.17%) | 0 (0%) | 1 (4.17%) |
| 129 | Telangiectasia | 1 (4.17%) | 1 (4.17%) | 0 (0%) |
| 130 | Tremor | 1 (4.17%) | 1 (4.17%) | 0 (0%) |
| 131 | Urinary retention | 1 (4.17%) | 0 (0%) | 1 (4.17%) |
| 132 | Urine output decreased | 1 (4.17%) | 0 (0%) | 1 (4.17%) |
| 133 | Wheezing | 1 (4.17%) | 1 (4.17%) | 0 (0%) |
