## Supplemental Table 2 for "Phase II study of nivolumab and ipilimumab for treatment of metastatic/recurrent adenoid cystic carcinoma (ACC) of all anatomic sites of origin and other malignant salivary gland tumors"

Supplementary Table 2: Molecular alterations among patients (n=4) with available NGS, RNA seq, and histochemical data

| Patient ID | Assay | TMB (m/MB) | PDL1 status | NGS findings (no VUSs) | Best response |
| --- | --- | --- | --- | --- | --- |
| 6 | Tissue NGS^a^ | 0.51 | Not available | BAP1 p.P293fs, MCL1 copy number gain, MDM4 copy number gain; BCOR p.L1130fs, NRAS copy number gain, PTGS2 copy number gain^25,26^ | PR |
| 7 | Tissue and ctDNA NGS^a^ | 1.1 on Tempus, Could not be calculated on Guardant | <1% | KDR copy number gain, PDGFRA copy number gain | SD |
| 10 | Tissue NGS^a^ | 0.4 | <1% | NOTCH1 p.C1383fs, MYB-NFIB chromosomal rearrangement; CKS1B copy number gain | PD |
| 14 | Tissue NGS and RNA expression analysis^a^ | 0.8 | Not available | ATM p.L2427R missense LOF, MYB-NFIB translocation; CUX1 c.564-1 G>A splice variant; FGFR2 and HRAS overexpressed | PD |

^a^Tissue NGS and RNA expression analysis from Tempus and ctDNA NGS from Guardant
