## Supplemental Figure 1 for "Phase II study of nivolumab and ipilimumab for treatment of metastatic/recurrent adenoid cystic carcinoma (ACC) of all anatomic sites of origin and other malignant salivary gland tumors"

A) RECIST v1.1 PFS and OS for combined patient cohort separated by neutrophil-lymphocyte ratio (NLR) of 5

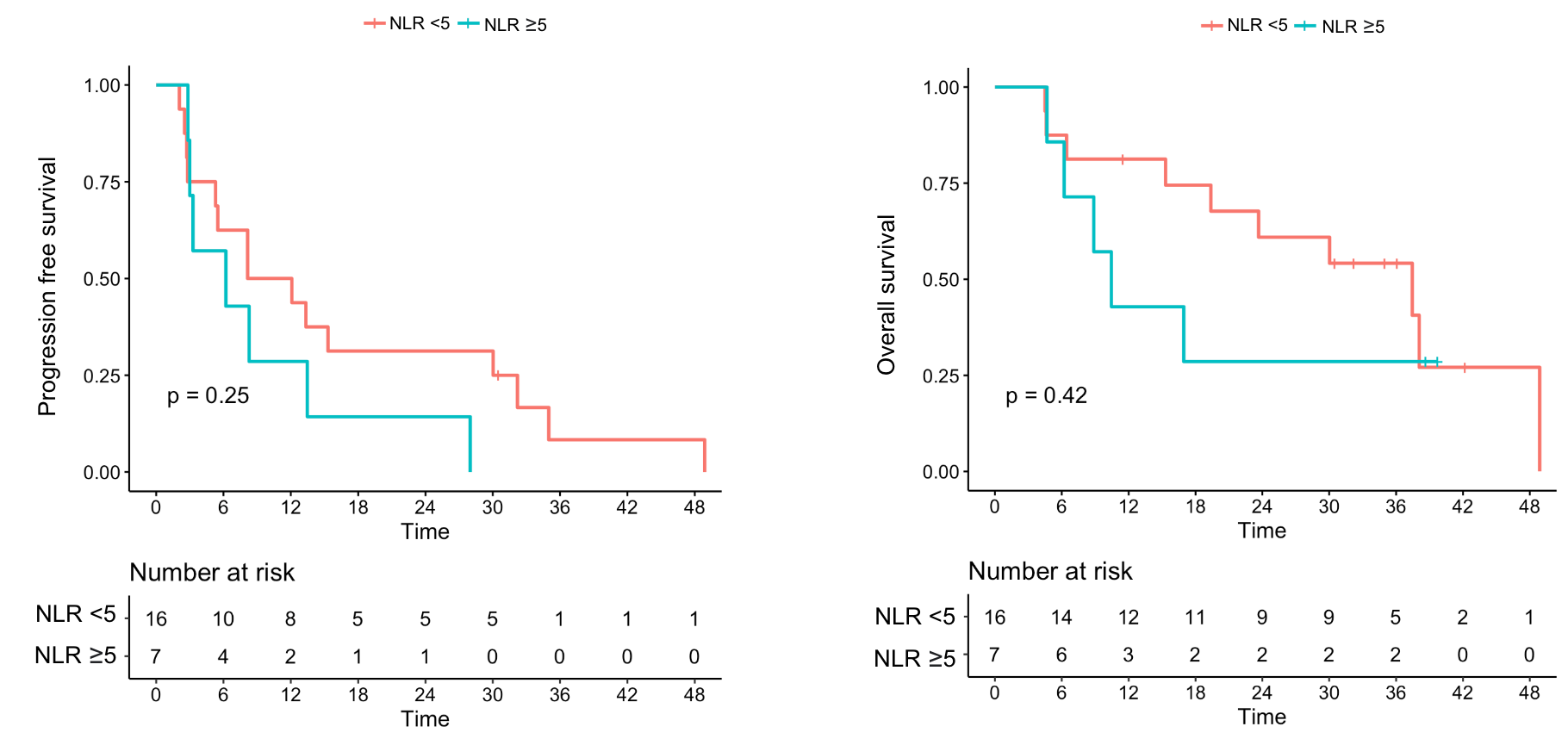

B) RECIST v1.1 PFS and OS for combined patient cohort separated by median neutrophil count

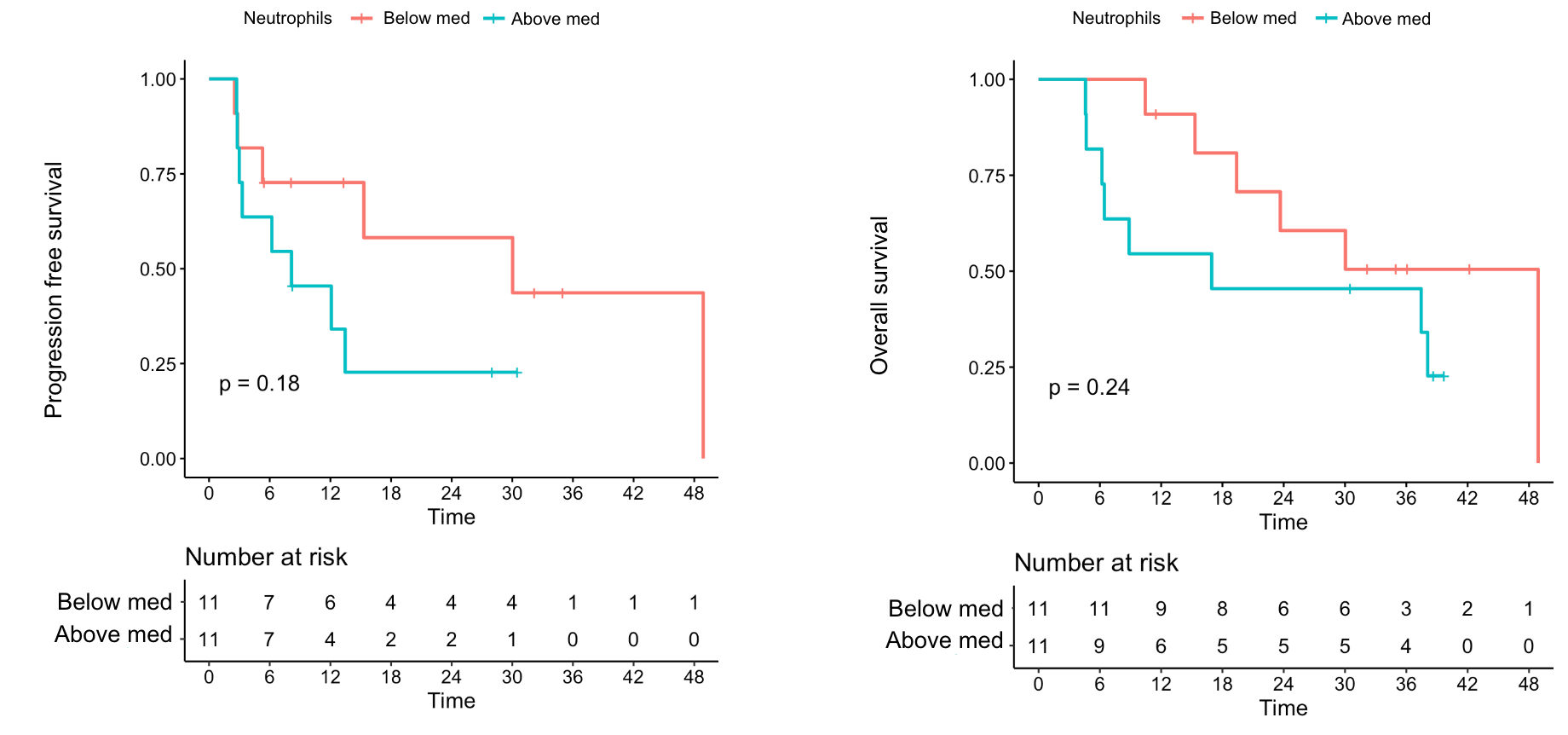

C) RECIST v1.1 PFS and OS for combined patient cohort separated by median platelet count

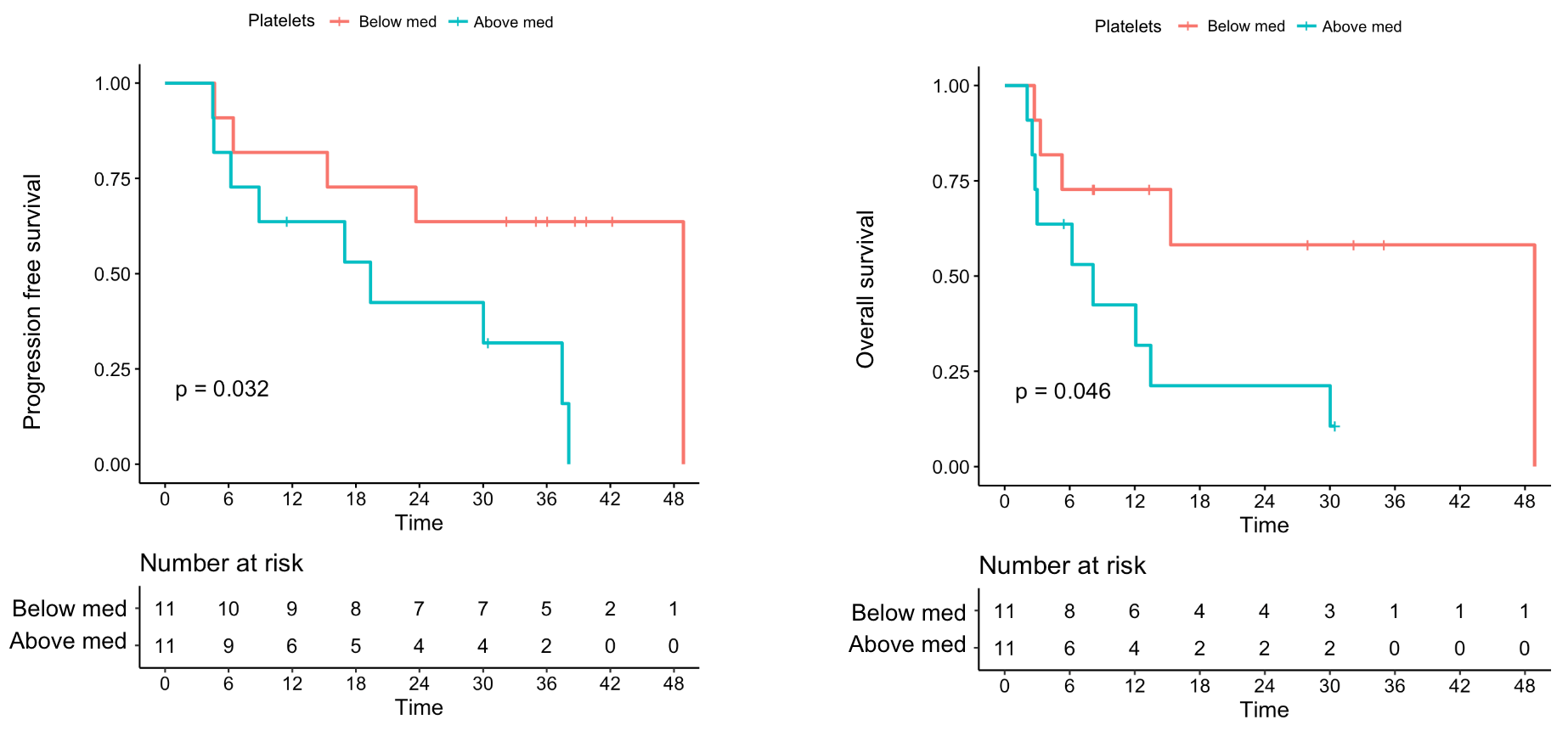

D) Pre-treatment predictors of outcomes

| **Variable** | **Measurement** | **Hazard Ratio** | **95% CI** |
| --- | --- | --- | --- |
| **Above-median platelet count** | PFS | 2.24 | 0.904-5.53 |
|  | OS | 3.60 | 1.11-11.7 |
| **Above-median neutrophil count** | PFS | 2.13 | 0.855-5.31 |
|  | OS | 2.14 | 0.709-6.47 |
| **Neutrophil-lymphocyte ratio ≥5** | PFS | 1.81 | 0.680-4.83 |
|  | OS | 1.70 | 0.547-5.28 |

*Median platelet count was 246K/uL

*Median neutrophil count was 4.3K/uL
